# Single-Blind and Double-Blind Peer Review: Effects on National Representation

**DOI:** 10.1101/2021.01.28.21250707

**Authors:** Meghana Kalavar, Arjun Watane, David Wu, Jayanth Sridhar, Prithvi Mruthyunjaya, Ravi Parikh

**Affiliations:** Bascom Palmer Eye Institute at the University of Miami Miller School of Medicine. 900 NW 17 Street. Miami, FL 33136; Wilmer Eye Institute, Johns Hopkins Hospitals Baltimore, MD; Department of Ophthalmology, Stanford University School of Medicine, Stanford, CA; Manhattan Retina and Eye Consultants New York, NY; New York University Langone Health New York, NY

**Author notes:** **Corresponding Author and Address for Reprints:** Ravi Parikh, Address: 67 E 78th Street Unit 1c, New York, NY 10075. Meeting Presentation: This data has not been submitted nor presented at any local or national conferences. Conflict of Interest: Dr. Sridhar is a consultant for Alcon, Dorc, Regeneron, and Oxurion. Dr. Parikh is a consultant for Anthem Blue Cross Blue Shield.

**Keywords:** peer review, disparity, masking

## Abstract

**Background/Objectives:** To assess whether the type of peer-review (single-blinded vs double-blinded) has an impact on nationality representation in journals.

**Methods:** A cross-sectional study analyzing the top ten nationalities contributing to the number of articles across 16 ophthalmology journals.

**Results:** There was no significant difference in the percentage of articles published from the journal’s country of origin between the top single-blind journals and top double-blind journals (SB= 42.0%, DB = 26.6%, p=0.49) but there was a significant difference between the percentage of articles from the US (SB=48.0%, DB=22.8%, p=0.02). However, there was no significant difference for both country of origin (SB =38.0%, DB =26.6%, p=0.43) and articles from the US (SB=35.0%, DB=22.8%, p=0.21) when assessing the top 8 double-blind journals matched with single-blind journals of a similar impact factor. The countries that most commonly made the top ten lists for highest number of articles were the US (n=16, 100%) and England (n=16, 100%). This held true even for journals established outside the United States (US=11/12, England=11/12).

**Conclusions:** There was no statistically significant difference in country-of-origin representation between single-blind journals and double-blind journals. However, higher income countries contributed most often to the journals studied even among journals based outside the US.

## Introduction

Several studies have noted a geographic disparity in the publication of manuscripts, with a greater proportion coming from countries with higher incomes.^1,2^ This is particularly relevant considering the growing number of article submissions from outside the United States.^3^ One study showed that global research output has increased annually by 4% between 2008 and 2018,^4^ while another found that international submissions rose from 9% in 1988 to 24% in 2000.^3^ Since many of the major peer-reviewed journals are established in high-income countries, one possible explanation is an associated geographical bias where reviewers of a certain country are more likely to accept articles from their own country or articles from nations with “track records.” ^2,5^ Studies have shown that US-based reviewers are more likely to accept US-based authors,^1^ and 36.4% of authors from lower-income countries (non-members of the Organization of Economic Co-operation and Development) believe there may be a bias against manuscripts published by authors from their own countries.^6^

Peer-review is critical as it provides the scientific community with confidence that there has been an examination of the scientific validity of the data and it is the gold-standard for evaluating work in scientific journals.^7^ While the majority of articles published in academic fields, including ophthalmology, are reviewed in a single-blinded manner, the ideal method of peer-review is often debated. Proponents of single-blind review (where reviewers have access to the authors’ names and nationality, but the authors are blinded to the reviewers) emphasize the high time/cost-burden associated with the double-blind peer review process and a lack of strong evidence that it is beneficial.^8,9^ This stands in contrast to a double-blinded review (where both reviewers and the authors are anonymized). In theory, a double-blinded approach ensures that any implicit biases the reviewer may hold with regards to gender, institution, geographic location, and ethnicity, etc. do not influence the decision to accept or reject a manuscript for publication. Proponents of a double-blinded review system state that single-blind reviewers are more likely to accept manuscripts published by well-known authors,^10,11^ creating a self-perpetuating cycle where well-known authors continue publishing and further increase their visibility.

Several studies across a variety of fields have attempted to quantify the impact of single-blind peer review vs double-blind peer review on the type of articles accepted. The effectiveness of masking from these papers have been mixed; while some state that masking does not impact the quality of peer review,^8,9^ others state that having a double-blinded peer review results in a higher number of acceptances for female authors,^12^ a higher rejection rate of well-known authors,^11,13^ and a higher number of authors from non-English speaking countries.^5,10^ However, to the best of the authors’ knowledge, no study has attempted to survey the peer review process in ophthalmology-related journals. Given the rise of international article submissions and the lack of concrete evidence on how reviewer behavior and submission acceptances differ based on the method of peer-review in ophthalmology, the goal of this study is to assess whether differences in national representation is dependent on the type of peer review process (single-blind vs double-blind) the journal has across 16 ophthalmology journals.

## Methods

We identified all the double-blind ophthalmic journals (of which there were 8) in the top 60 journals by impact factor as reported by the Institute of Scientific Information’s annual Journal Citation Reports and compared them to the top 8 single-blinded journals on the list.^14^ Journals were determined to be double-blind or single-blind either through the journal website, the journal submission site, or by emailing the editorial office. Journals that were invitation-only were excluded. Furthermore, we performed additional analysis on 8 single-blind journals that were matched by impact factor to the top 8 double-blind journals. The Journal of Citation Reports was also used to determine the top ten nationalities that contributed to published articles in each journal between 2017 and 2019. If there was a tie for tenth place amongst various nationalities, only the first alphabetically was included (so every journal only had ten countries total). The Mann Whitney U Test was used to compare medians between single-blind journals and double-blind journals.

## Results

### Top 8 Double-Blind vs Top 8 Single-Blind by Impact Factor

Eight out of the 16 highest impact factor single-blinded/double-blind journals were based out of the United States. The average impact factor of the top eight single-blind journals was 5.74, and the average impact factor for double-blind journals was 1.55 (p=0.001). The top single-blinded journals were most commonly based in the US (6/8) whereas the top double-blinded journals were most commonly based in the Netherlands (3/8). The countries that most commonly made the top ten lists for highest number of articles were the US (n=16, 100%), England (n=16, 100%), and China (n=15, 94%). Of the 32,081 articles included in this analysis, 46.2% were from the US, 8.5% were from China, and 7.5% were from England. There was no significant difference in the percentage of articles published from the journal’s country of origin between the top single-blind journals and top double-blind journals (SB= 42.0%, DB = 26.6%, p=0.49) but there was a significant difference between the percentage of articles from the US (SB=48.0%, DB=22.8%, p=0.02) (Table 1).

**Table 1:**

| Top Double-Blind Journals |  |  |  |  | Top Single-Blind Journals |  |  |  |  | Matched Single-Blind Journals |  |  |  |  |
| --- | --- | --- | --- | --- | --- | --- | --- | --- | --- | --- | --- | --- | --- | --- |
| Name | Impact Factor | Country of Origin | % Articles from Country of Origin | % Articles from US | Name | Impact Factor | Country of Origin | % Articles from Country of Origin | % Articles from US | Name | Impact Factor | Country of Origin | % Articles from Country of Origin | % Articles from US |
| Contact Lens and Anterior Eye | 2.578 | Netherlands | 0.00% | 17.10% | Ocular Surface | 12.336 | Netherlands | 0.00% | 42.00% | JoN | 2.513 | US | 66.20% | 66.20% |
| CEO | 1.918 | US | 6.90% | 6.90% | Ophthalmology | 8.47 | US | 52.70% | 52.70% | Current Eye Research | 1.754 | US | 21.00% | 21.00% |
| Cutaneous and Ocular Toxicology | 1.385 | England | 1.50% | 13.80% | JAMA | 6.198 | US | 68.90% | 66.40% | CJO | 1.369 | Canada | 47.80% | 23.80% |
| JAAPOS | 1.339 | US | 64.20% | 62.50% | SoO | 4.195 | US | 51.60% | 54.20% | OPRS | 1.331 | US | 62.40% | 62.40% |
| IJO | 1.33 | China | 58.30% | 10.20% | AJO | 4.013 | US | 53.20% | 53.20% | IO | 1.31 | Netherlands | 0.00% | 9.20% |
| Ophthalmic Genetics | 1.308 | Netherlands | 0.00% | 37.50% | Retina | 3.649 | US | 37.10% | 37.10% | Perception | 1.217 | England | 22.50% | 13.80% |
| DO | 1.294 | Netherlands | 0.00% | 28.30% | BJO | 3.611 | England | 18.10% | 24.40% | SIO | 1.205 | US | 34.30% | 34.30% |
| Indian Journal of Ophthalmology | 1.25 | India | 82.10% | 6.10% | IOVS | 3.47 | US | 54.30% | 54.30% | JPOS | 1.1 | US | 49.70% | 49.70% |
| Average | 1.55 |  | 26.60% | 22.80% |  | 5.74 |  | 42.00% | 48.00% |  | 1.47 |  | 38.00% | 35.00% |
|  |  |  |  |  | P-Value comparing Top Double-Blind Journals with Top Single-Blind Journals | 0.001 |  | 0.49 | 0.02 | P-Value comparing Top Double-Blind Journals with Matched Single-Blind Journals by Impact Factor | 0.46 |  | 0.43 | 0.21 |

CEO = Clinical and Experimental Ophthalmology, JAAPOS = Journal of AAPOS, IJO = International Journal of Ophthalmology, DO = Documenta Ophthalmologica, JAMA = JAMA Ophthalmology, SoO = Survey of Ophthalmology, AJO = American Journal of Ophthalmology, Retina = Retina - The Journal of Retinal and Vitreous Diseases, BJO = British Journal of Ophthalmology, IOVS = Investigative Ophthalmology & Visual Science, CJO = Canadian Journal of Ophthalmology, OPRS = Ophthalmic Plastic and Reconstructive Surgery, IO = International Ophthalmology, JoN = Journal of Neuro-ophthalmology, SIO – Seminars in Ophthalmology, JPOS = Journal of Pediatric Ophthalmology and Strabismus
\*P-Values are calculated by the Wilcoxon Rank-Sum Test. Values less than 0.05 are considered significant.

### Top 8 Double-Blind Matched with Top 8 Single-Blind by Impact Factor

As there was a significant difference in impact factor between the top double-blind and top single-blind studies, we matched the top double-blind studies with single-blind studies by impact factor. In this analysis, there was no significant difference in impact factor between single-blind and double-blind journals (SB=1.47, DB=1.55, p=.46). There was no significant difference between the top 8 double-blind journals matched with single-blind journals of a similar impact factor for both country of origin (SB =38.0%, DB =26.6%, p=0.43) and articles from the US (SB=35.0%, DB=22.8%, p=0.21) (Table 1).

### Top 6 Double-Blind International Journals Matched with Single-Blind International Journals of Similar Impact Factor

Amongst journals that were not based in the US, we matched journals by impact factor to see whether there was any difference in US representation amongst international journals (Table 2). The average impact factor of double-blind international journals was 1.52 and the average impact factor for matched single-blind international journals was 1.31 (p=0.42). There was no difference between single-blind and double-blind journals regarding both country of origin (SB=38.3%, DB= 23.8%, p=0.37) and US representation (SB=14.1%, DB=18.8%, p=0.38). Of the journals that were based outside the US, England (n=11/12) and the US (n=11/12) most frequently occurred in the top ten nations that contributed articles. India contributed the majority of articles (n=1,570, 25.2%) followed by the US (n=1,088, 17.5%) and Germany (n=1,028, 16.5%).

**Table 2:**
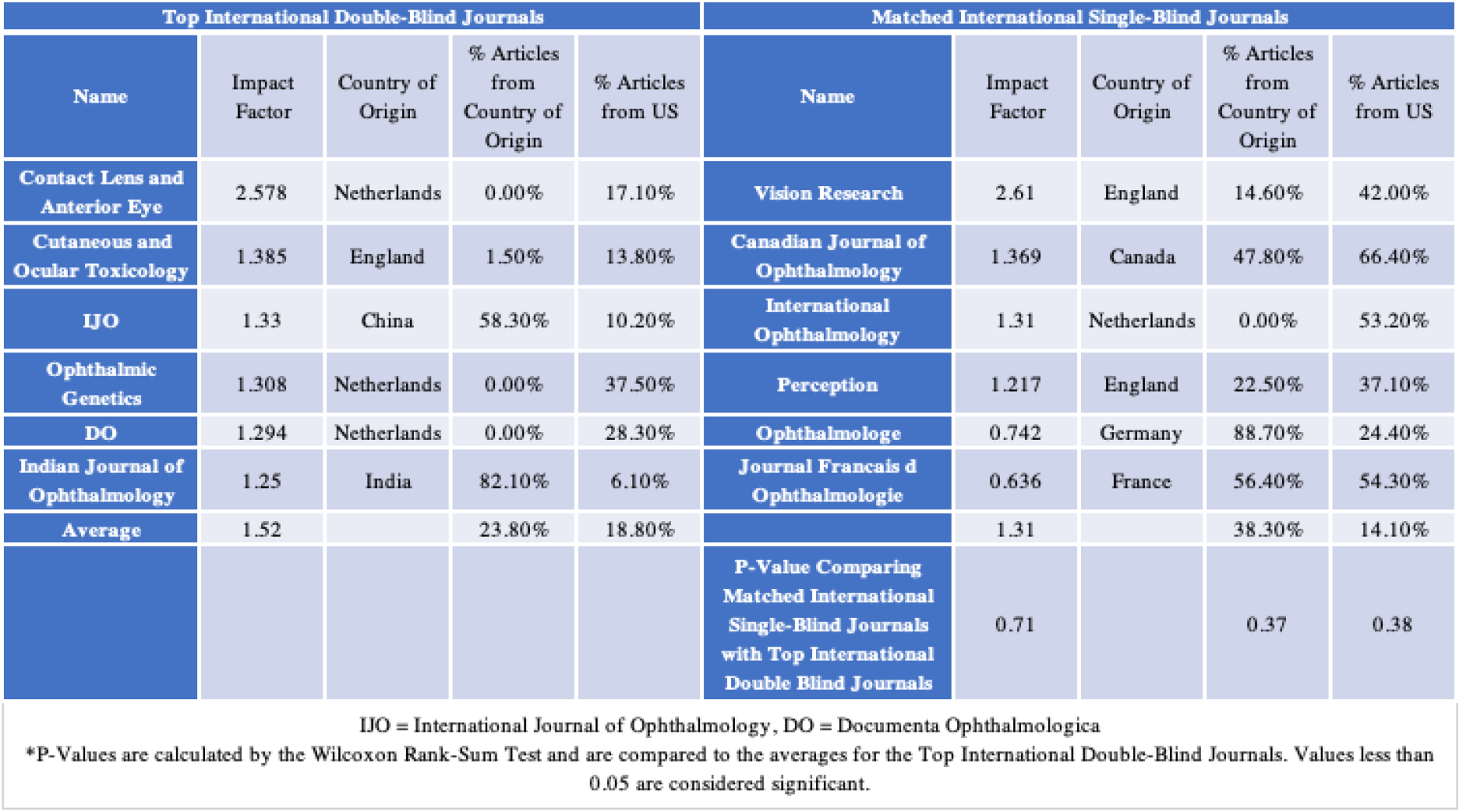

| Top International Double-Blind Journals |  |  |  |  | Matched International Single-Blind Journals |  |  |  |  |
| --- | --- | --- | --- | --- | --- | --- | --- | --- | --- |
| Name | Impact Factor | Country of Origin | % Articles from Country of Origin | % Articles from US | Name | Impact Factor | Country of Origin | % Articles from Country of Origin | % Articles from US |
| Contact Lens and Anterior Eye | 2.578 | Netherlands | 0.00% | 17.10% | Vision Research | 2.61 | England | 14.60% | 42.00% |
| Cutaneous and Ocular Toxicology | 1.385 | England | 1.50% | 13.80% | Canadian Journal of Ophthalmology | 1.369 | Canada | 47.80% | 66.40% |
| IJO | 1.33 | China | 58.30% | 10.20% | International Ophthalmology | 1.31 | Netherlands | 0.00% | 53.20% |
| Ophthalmic Genetics | 1.308 | Netherlands | 0.00% | 37.50% | Perception | 1.217 | England | 22.50% | 37.10% |
| DO | 1.294 | Netherlands | 0.00% | 28.30% | Ophthalmologe | 0.742 | Germany | 88.70% | 24.40% |
| Indian Journal of Ophthalmology | 1.25 | India | 82.10% | 6.10% | Journal Francais d Ophthalmologie | 0.636 | France | 56.40% | 54.30% |
| Average | 1.52 |  | 23.80% | 18.80% |  | 1.31 |  | 38.30% | 14.10% |
|  |  |  |  |  | P-Value Comparing Matched International Single-Blind Journals with Top International Double Blind Journals | 0.71 |  | 0.37 | 0.38 |

IJO = International Journal of Ophthalmology, DO = Documenta Ophthalmologica
\*P-Values are calculated by the Wilcoxon Rank-Sum Test and are compared to the averages for the Top International Double-Blind Journals. Values less than 0.05 are considered significant.

## Discussion

Our results demonstrate there was no difference in the number of articles published from the journal’s country of origin between single-blind journals and double-blind journals. These results are promising given the importance of publishing in peer-review journals on an international stage, both for the dissemination and creation of knowledge but also for academic advancement. Of note, we did find that the top single-blind journals had a greater percentage of journals from the US, but we believe this is due to the large number of high-impact factor, single-blind journals based in the US. In order to eliminate this confounding, we matched the top double-blind journals by impact factor to single-blind journals and found that this difference was eliminated. These results are in line with another study looking at abstracts submitted to the American Heart Association conference which found that double-blind review helps mitigate geographic disparities as significantly fewer abstracts were accepted from the US during double blind review.^5,10^

A previous study demonstrated that while both US and non-US based authors view “non-US” studies similarly, US reviewers have a significant preference for US authors.^1^ However, our study found that single-blind journals and double-blind journals, when matched by impact factor, had similar rates of article publication from the US. This is congruent with conclusions from other studies describing increased representation of high-income countries in journals.^2,5^ Furthermore, we found that even when considering only journals that were based outside of the US, England and the US (both high-income countries) most commonly made the top-ten lists for contributing countries. These findings support another study that determined 80% of published biomedical articles originate from ten countries.^15^ Of note, the findings of our study may be confounded given that the populations submitting to the articles may be different depending on the scope of the journal. However, a study evaluating submissions from authors who had the option to elect single-blind or double-blind review in *Nature* journals found that articles under double-blind review tended to get rejected more often. The authors of the aforementioned study attributed the higher rejection rate of double-blind review to poorer quality of the articles.^16^ High-income countries are more likely to have increased institutional funding,^17^ increased cross-collaboration with other high-income countries,^18^ and prioritization of research by the institution, all of which positively impact the quality of a manuscript. For similar reasons, it may also be that lower-income countries tend to have lower research output. Furthermore, another study found that poor writing was one of the top six reasons reviewers reject a manuscript.^3^ Considering the majority of high-impact journals are in English, as was corroborated in this study, many countries whose native-language is not English may be at an inherent disadvantage.^3^ Further studies investigating potential interventions to improve the quality of manuscripts from international countries to increase the chances of publishing in these high-impact journals are necessary.

Editors have long been a proponent of single-blind review. One commonly cited reason for this is the importance of knowing the author’s identity to highlight strengths and weaknesses of the work. For example, having a statistician as a co-author on a manuscript could further validate the statistical methods of a paper. Similarly, knowing the identity of the author could be important in order to distinguish progress from previous works.^19^ Another commonly cited reason for masking reviewers is an overwhelming burden on the journal to ensure that true masking is achieved. This is due to reviewers being able to commonly guess the authors, time and labor costs required to educate the reviewers, and issues regarding reconciling the blind with the unblinded manuscript. In fact, one of the biggest issues with double-blind review is the issue of true masking. Often, the reviewer can infer the author, particularly if the author is well-known in the field, due to references to study setting, pronouns used in the manuscript, or the author’s previous works. One study in economic journals showed that 45% of reviewers can identify the author,^20^ while another study found masking failure to be 32%.^21^ In ophthalmology, specifically, this can be an even greater issue given the relatively small size of the field. One suggested remedy for this is to use reviewers that have less research and reviewer experience, but this may have unintended consequences regarding the quality of review.^22^ Our study appears to validate further the use of single blind review among the top journals in the field. However, additional studies evaluating other sources of potential bias are necessary.^12^

This study has limitations. First, while the journals are located in one country, the national representation of the editorial boards/reviewers editing them could be from different countries which may affect article acceptance. Second, this study does not account for article submission rates or researcher population rates of countries as that data is not publicly available. Both of these factors could significantly influence the number of articles published in a journal. We were also not able to take into account the reach of the journal; while some journals may be known on an international level and thus receive submissions from across the world, other journals may only be more prominent locally thus warranting more submissions from the nation where that journal is based.

While our study found that double blind review did not impact the number of articles accepted from outside the country of origin, our findings were consistent with other studies demonstrating a predominance of articles from higher-income countries. This study evaluates one component of potential disparities and implicit bias in the peer-review process. Further studies assessing the role of peer-review on other possible disparities (i.e. prestige or gender of author) in ophthalmology journals are necessary. Furthermore, additional studies taking into account submission and acceptance rates in single-blind vs double-blind scenarios are warranted to more accurately quantify the existence of geographical disparities so that we can ensure a more equitable publishing environment.

## Data Availability

All data was collected from the Journal Citation Reports produced by the Institute of Scientific Information

